# Cohort profile: The *SmartSleep Study*, Denmark Triangulation of evidence from survey, clinical and tracking data

**DOI:** 10.1101/2022.12.19.22283650

**Authors:** NH Rod, TO Andersen, ER Severinsen, C Sejling, AS Dissing, VT Pham, M Nygaard, LKH Schmidt, HJ Drews, TV Varga, NlC Freiesleben, HS Nielsen, AK Jensen

**Affiliations:** Section of Epidemiology, Department of Public Health, University of Copenhagen, Denmark; Department of Obstetrics and Gynecology, Copenhagen University Hospital Hvidovre, Denmark; Section of Biostatistics, Department of Public Health, University of Copenhagen, Denmark

## Abstract

**Purpose:** The *SmartSleep Study* is established to comprehensively assess the impact of night-time smartphone use on sleep patterns and health. An innovative combination of large-scale repeated survey information, high-resolution sensor-driven smartphone data, in-depth clinical examination and registry linkage allow for detailed investigations into multisystem physiological dysregulation and long-term health consequences associated with night-time smartphone use and sleep impairment.

**Participants:** The *SmartSleep Study* consists of three interconnected data samples, which combined include 30,673 individuals with information on smartphone use, sleep and health. Subsamples of the study population also include high-resolution tracking data (n=5,927) collected via a customized app and deep clinical phenotypic data (n=245). A total of 7,208 participants will be followed in nationwide health registries with full data coverage and long-term follow-up.

**Findings to date:** We highlight previous findings on the relation between smartphone use and sleep in the *SmartSleep Study*, and we evaluate the interventional potential of the citizen science approach used in one of the data samples. We also present new results from an analysis in which we utilize 803,000 data-points from the high-resolution tracking data to identify clusters of temporal trajectories of night-time smartphone use that characterize distinct use patterns. Based on these objective tracking data, we characterize four clusters of night-time smartphone use.

**Future plans:** The unprecedented size and coverage of the *SmartSleep Study* allow for a comprehensive documentation of smartphone activity during the entire sleep span. The study will be expanded by linkage to nationwide registers, which will allow for further investigations into the long-term health and social consequences of night-time smartphone use. We also plan new rounds of data collection in the coming years.

**STRENGTHS and LIMITATIONS of this study:**

- The unprecedented size and coverage of the *SmartSleep Study* allow for a comprehensive objective and subjective documentation of smartphone activity during the entire sleep span.
- The data in the *SmartSleep Study* are sampled by three different strategies, which allow us to test robustness and validate findings across samples. This aligns with the principles of triangulation, which aims at obtaining more reliable answers to complex research questions through the integration of results from different approaches with different sources of bias.
- The *SmartSleep Study* is readily available for research projects: the data sources have already been linked, the data have been cleaned and prepared for future analyses.
- The *SmartSleep Study* is not fully representative of the general population due to the sampling procedures, and we are currently creating weights that can be used in the statistical analysis to compensate for this imbalance.

## INTRODUCTION

Sleep facilitates central biological processes involving restitution and brain plasticity.^1^ Disruption of these essential processes may lead to severe health consequences.^2^ Smartphones are easily carried into bed and offer multiple facilities that may disrupt sleep through mental activation (e.g., social media, streaming, news feeds). Also, the artificial light exposure from smartphones and the emission of electromagnetic radiation can delay release of sleep hormones and affect sleep latency.^3–5^ Thus, the massive and increasing 24-hour use of smartphones raises public health concern. The *SmartSleep Study* is established to comprehensively assess the impact on night-time smartphone use on sleep patterns and health. An innovative combination of large-scale repeated survey information, high-resolution sensor-driven smartphone data, in-depth clinical examination and registry linkage allow for detailed investigations into multisystem physiological dysregulation, obesity, sub-optimal mental health, fertility, and long-term health consequences associated with night-time smartphone use and sleep impairment.

The rising prevalence of mental health problems in young adulthood is alarming. While sleep problems are well-known symptoms of depression, sleep disturbances have also been shown to play an etiological role in the development of mental illness through altered emotion regulation and neurobiological interaction.^6,7^ Sleep is often perceived to be a modifiable risk factor, but to restore good sleep we need to address the problem of unhealthy sleep habits. Recent reviews show a clear correlation between mobile phone addiction and anxiety, depression, and sleep quality.^8,9^ This highlights a need for large-scale studies documenting the effect of different types of night-time smartphone behavior on mental health outcomes. Linkage of high-resolution smartphone tracking data with detailed health registries in the *SmartSleep Study* allows for documentation of the dynamic interplay between night-time smartphone use, sleep, and mental health problems.

Infertility is another issue of public health concern. Only one in four young men has optimal semen quality,^10^ and one week restriction of sleep to 5 hours per night was related to lower serum testosterone levels in an experimental study.^11^ It has also previously been shown that young men with self-reported disturbed sleep have lower semen quality than men with less disturbed sleep in a general population sample.^12^ While the majority of studies on risk factors for dysfunctioning of the reproductive system are based on infertile male populations, the *SmartSleep Study* includes a detailed clinical examination of irregularities of menstrual cycles and ovulation in young women before they are aware of their own fertility potential.

Previous studies have reported negative effects of technology use on sleep, health, and well-being,^9,13–15^ but the validity and translation of these findings are limited by the fact that they are mainly cross-sectional in design, the majority are based on self-reported smartphone use and restricted to smartphone use around bedtime primarily among children, adolescents, and young adults. Only a few smaller studies have objectively measured screen time in relation to health outcomes.^16,17^ The *SmartSleep Study* provides an unprecedented data infrastructure that will add important new insights into the health consequences of smartphone interrupted sleep, which goes well beyond traditional epidemiological studies based on single questionnaire-based measures of sleep.

## STUDY DESCRIPTION

### The study population

The *SmartSleep Study* consists of three interconnected data samples which ensure an innovative combination of large-scale repeated survey information, high-resolution sensor-driven smartphone data, and in-depth clinical examination. A flowchart outlines the participants in each data sample (Figure 1), and the timeline for the data collection is presented in Suppl Figure 1.

**Figure 1.**
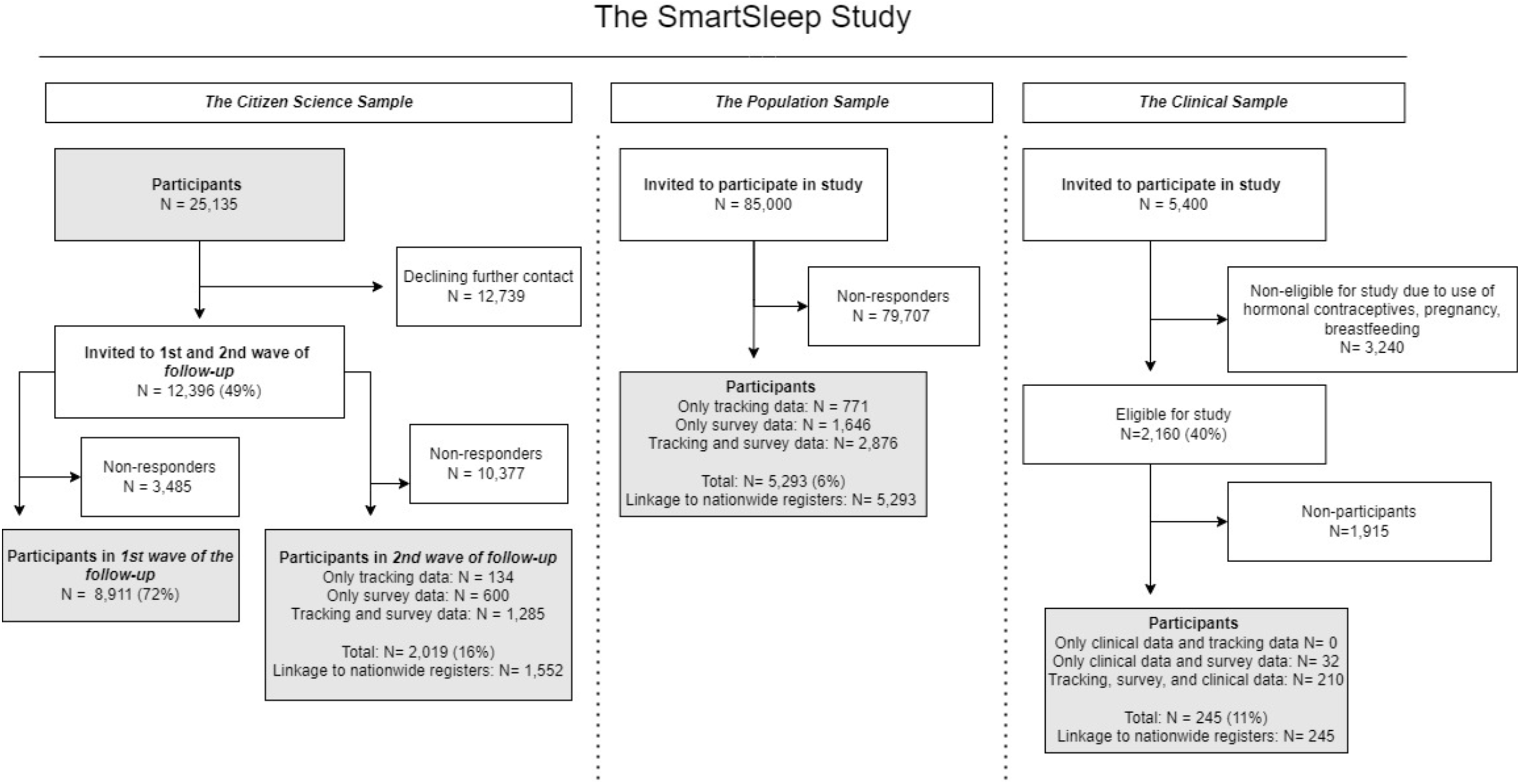
Flow chart for the data collection of the *SmartSleep Study*

#### The Citizen Science Sample

The *baseline data collection* of the Citizen Science Sample included 25,135 Danish adults, who were recruited in November 2018 using a citizen science approach. The Danish National Broadcasting Corporation (DR) was used as a vehicle for creating attention and public interaction around the research project and formed the framework for recruiting the study participants. This approach was inspired by other similar citizen science studies, such as the British Broadcasting Corporation (BBC) Loneliness Experiment^18^ and the Rest Test.^19^ Citizen science more generally aims to benefit both researchers and the participants by transferring knowledge between each other; the researchers obtain data for research, and the participants gain literacy and knowledge on scientific topics.^20^

During the week of data collection, the DR deployed a theme about sleep on several of their national radio programs, their website, and social media pages. The theme was intended as a wrapper of the *SmartSleep* survey and to engage participants in discussions concerning sleep and smartphone use. To create a feedback process between researchers and participants, preliminary results were generated daily and reported on the <u>project website</u>. The preliminary results were also reported live on the radio followed by interviews with the researchers. A personal feedback feature was incorporated in the survey, providing each participant with individual, direct feedback on their reported night-time smartphone use compared to an age- and gender-stratified mean. The feedback was intended to make the participants reflect on their own sleep and smartphone behavior. Participants had to be aged 16 years or older and understand written Danish to participate. A detailed description of data collection for the Citizen Science Sample has previously been published.^21^

*The 1*^*st*^ *wave of follow-up* was initiated already two weeks after the citizen science data collection had ended (November/December 2018). A total of 12,396 participants in the baseline data collection (49%) had indicated that we could contact them for future studies. These people received an online follow-up survey via email. The follow-up survey was very short and aimed at evaluating whether the participants had changed their smartphone behavior following the citizen science campaign. Thus, the 1^st^ wave follow-up survey only focused on night-time smartphone use, changes in night-time smartphone use, and motivational factors for these changes. Up to three reminders were sent to those not responding to the first email. In total, 8,911 responded to the follow-up survey (72% response rate). See Andersen et al.^22^ for more details on this data collection.

*The 2*^*nd*^ *wave of follow-up* data was collected between February and July 2020, where the 12,396 participants who volunteered for future studies were invited via email to participate in a second follow-up. The median follow-up period from the baseline data collection was 543 days, corresponding to 18 months (IQR: 530-555 days). This wave of data collection was rather extensive and included a) filling in a detailed survey, and b) downloading the *SmartSleep* app and tracking one’s night-time smartphone use for up to 14 nights. A total of 1,426 participants provided both tracking and survey information, 459 provided only survey data, and 164 provided only tracking data (17% response rate). The participants in the Citizen Science Sample on average tracked 10.5 (SD: 13.3) nights/median 7 nights (IQR: 2-14).

#### The Population Sample

The Citizen Science Sample was supplemented with a random population sample to ensure a broader representation of the general population. We invited 85,000 Danish adults between 18-50 years of age between July and October 2020 randomly selected from the Central Population Register. The participants were invited via a secure digital postbox which was identified via the participants’ social security number. The same methodology for data collection as in the 2^nd^ wave follow-up of the Citizen Science Sample was followed; the participants were asked to fill in a detailed survey, as well as download the *SmartSleep* app and track their night-time smartphone use up to 14 nights. Up to two reminders were sent to non-responders. In total, 3,222 participants provided both survey and tracking information, 1,300 only provided survey information, and 889 only provided tracking data (6% response proportion). The participants in the Population Sample on average tracked 5.3 (SD: 4.7) nights/median 4 nights (IQR: 2-8).

#### The Clinical Sample

The Clinical Sample was aimed at obtaining clinical and biological measures to investigate whether night-time smartphone use associates with early signs of physiological dysfunctioning across multiple biological systems among young women. We invited 5,400 randomly selected women aged 18-25 years and living in proximity to the examination site of Copenhagen University Hospital Hvidovre in March 2019 from the Central Population Register. The data collection took place between November 2019 and October 2020 and was paused mid-April to end-June 2020 due to the outbreak of COVID-19 in Denmark. The women were invited via a secure digital postbox identified by their social security number. Pregnant women, breastfeeding women, and women using hormonal contraception were not eligible for the study, neither were women who did not have sufficient understanding of Danish. Exclusion criteria were clearly stated in the invitation letter and based on national statistics on pregnancy and use of contraceptives, we expected that approximately 40% (n∼2,160) of the invited random sample would be in fact eligible to participate. Among the eligible women, 245 women signed an informed consent and went through a comprehensive clinical examination (11% response proportion). The participants were also asked to download the *SmartSleep* app and track their night-time smartphone use for at least 7 nights and respond to a detailed survey. Of the 245 women participating in the clinical examination, 224 provided both survey and tracking data and 18 provided only survey data and two provided only tracking data. We offered DKK 500 (approx. € 67) and the possibility to receive personal test results in compensation for the participants’ time and willingness to participate. The participants in the Clinical Sample on average tracked 9.1 (SD: 6.2) nights/median 8 nights (IQR: 5-12).

*The Clinical Sample* is restricted to young women, but fertility issues in young men associated with night-time smartphone use and disturbed sleep are of course of equal importance. Thus, we have initiated a collaboration with an ongoing study on male fertility, in which we have included most of the smartphone and sleep-related survey items from the *SmartSleep Study*, starting from 2020 and onwards. This study annually enrolls approx. 300 young men into a comprehensive study on fertility status and semen quality. For details about the male fertility measures please see Priskorn et al.^23^

### Data types

#### Survey data

Survey data was collected in all samples. Table 1 provides an overview of the different topics covered in the surveys and in the different samples. We used items from the following validated scales: the Copenhagen Social Relations Questionnaire^24^, the sleep quality dimension of the Karolinska Sleep Questionnaire^25^, the 10-item Perceived Stress Scale^26,27^, the Major Depressive Inventory^28^, Satisfaction with Life Scale^29^, and the UCLA Loneliness short three-item T-ILS version^30^. The survey also included several items developed specifically for the study evaluating nightly smartphone behaviors. Smartphone dependency was measured using a subscale of the validated Problematic Mobile Phone Use Questionnaire^31^. In order to make use of the scale in Danish, we translated and back-translated the scale using authorized translators and subject matter experts. The back-translated version was reviewed independently by researchers and adjusted accordingly.

**Table 1.** Overview of survey data, clinical measures, biomarkers, and possibilities for long-term registry-linkage follow-up in the *SmartSleep Study*.

|  |  | Citizen science sample |  |  | Population Sample | Clinical Sample |
| --- | --- | --- | --- | --- | --- | --- |
|  |  | Baseline | 1<sup>st</sup> round | 2<sup>nd</sup> round | Baseline | Baseline |
|  | Populations, n | 25,135 | 8,911 | 2,019 | 5,293 | 245 |
| **Survey data** | Socio-demographics | x |  | x | x | x |
|  | Daytime smartphone use | x |  | x | x | x |
|  | Night-time smartphone use | x | x | x | x | x |
|  | Changes in smartphone use |  | x |  |  |  |
|  | Motivation for changes |  | x |  |  |  |
|  | Social media use | x |  | x | x | x |
|  | Smartphone dependency | x |  | x | x | x |
|  | Sleep, quantity and quality | x |  | x | x | x |
|  | Night work |  |  | x | x | x |
|  | Social relations |  |  | x | x | x |
|  | Self-reported health | x |  | x | x | x |
|  | Self-reported height/weight | x |  | x | x | x |
|  | Headache | x |  | x | x | x |
|  | Symptoms |  |  | x | x | x |
|  | Previous diagnoses |  |  | x | x | x |
|  | Common infections |  |  | x | x | x |
|  | Fertility and reproductive health |  |  | x | x | x |
|  | Sexual health |  |  | x | x | x |
|  | Contraceptives |  |  | x | x | x |
|  | Perceived stress | x |  | x | x | x |
|  | Depressive symptoms |  |  | x | x | x |
|  | Life satisfaction |  |  | x | x | x |
|  | Loneliness |  |  | x | x | x |
|  | Smoking |  |  | x | x | x |
|  | Alcohol consumption |  |  | x | x | x |
|  | Physical activity |  |  | x | x | x |
|  | Diet |  |  | x | x | x |
| **Clinical measures** | Weight/height |  |  |  |  | x |
|  | Hip/waist circumference |  |  |  |  | x |
|  | Blood pressure |  |  |  |  | x |
|  | Resting pulse |  |  |  |  | x |
|  | Ovarian morphology |  |  |  |  | x |
|  | Uterine morphology |  |  |  |  | x |
|  | Antral follicle count (AFC) |  |  |  |  | x |
|  | Volume of the ovaries |  |  |  |  | x |
|  | Menstrual cycle (interview) |  |  |  |  | x |
| **Biomarkers** | Total cholesterol |  |  |  |  | x |
|  | VLDL cholesterol |  |  |  |  | x |
|  | LDL cholesterol |  |  |  |  | x |
|  | HDL cholesterol |  |  |  |  | x |
|  | Triglycerides |  |  |  |  | x |
|  | HbA1c |  |  |  |  | x |
|  | Glucose |  |  |  |  | x |
|  | Vitamin D |  |  |  |  | x |
|  | Immune system and inflammatory profiles |  |  |  |  | x |
| **Registry linkage follow-up** | Hospital diagnoses |  |  | x | x | x |
|  | Mortality |  |  | x | x | x |
|  | Prescription medication |  |  | x | x | x |

#### Tracking data

We developed an app (the *SmartSleep* app), which allowed us to continuously track all screen activations from participants during the sleep period. To increase the usability of the app, it was developed both for iOS and Android systems and it could function with multiple (newer) software versions on both systems. A dynamic panel was built into the app where participants could enter their daily sleep onset and offset times. A feedback panel was also made available in the app where the participants could follow their own screen time use during the sleep period. This panel was added to motivate the participants to use the app, but also to provide the participants with concrete examples of what type of information was collected and what they could be used for. The app could be downloaded either from App Store or from a provided link. Each participant was given a unique user-id in the invitation letter, which was used for entering the *SmartSleep* app. This user-id was used to link the different types of data. The *SmartSleep* app was tested for technical issues on several of the most used iOS and Androids models in Denmark. User experience was tested among ten voluntary test persons who were interviewed after having used the app for at least one week. For a small subsample of the health examination participants using the *SmartSleep* app iOS version (N=28), the collected screen activation data were compared to iOS’s own built-in screen data recordings, and reasonable overlap between the two was found (spearman correlation, r=0.72, please see Supplementary Text 1), although the *SmartSleep* app tended to record more activity than the iOS’s own built-in screen data recordings. Which one provide the most correct measure cannot be determined from this comparison. The app was built as open source and all available code can be accessed via GitHub: https://github.com/smartsleep/about

#### Clinical data and biomarkers

An overview of clinical data and biomarkers can be seen in Table 1. All clinical measures and biomarkers were collected at Copenhagen University Hospital Hvidovre as part of a comprehensive health examination of the 245 women in the *Clinical Sample*. The health examination consisted of measurement of weight, height, waist circumference, hip circumference, resting pulse and blood pressure. Further, non-fasting blood samples were collected (n=241) and the participants were evaluated by transvaginal ultrasonography (n=232) or transabdominal ultrasonography (n=13) to collect information on ovarian morphology and any uterine pathology, the number of ovarian follicles (antral follicle count [AFC]) and the volume of both ovaries to investigate indicators of reproductive function. Ultrasonography was performed during the early/mid follicular phase of the menstrual cycle (median of cycle day 7 (IQR:5-9)). In addition, the woman’s menstrual cycle length since menarche was systematically reviewed by interview with a focus on the last 3-6 months. Plasma was sampled and analyzed for levels of vitamin D, total cholesterol, triglyceride, high-density lipoprotein (HDL) cholesterol, very-low-density lipoprotein (VLDL) cholesterol, and low-density lipoprotein (LDL) cholesterol, which were analyzed sampled in Li-heparin tubes with a separation gel, and glycosylated hemoglobin (HbA1c), which was sampled in K2-EDTA tubes. These plasma analyses were processed at the same laboratory at the Department of Clinical Biochemistry at Copenhagen University Hospital Hvidovre (Cobas 8000, TOSOH HLC – 723 G8). Details about each procedure can be found at the Department of Clinical Biochemistry <u>website</u>. Immune system and inflammatory profiles were measured using a high-profile panel of 54 different human biomarkers including proinflammatory markers, cytokines and chemokines among others by a commercially available multiplexed immunoassay (V-PLEX Human Biomarker 54-plex) from Meso Scale Discovery (Rockville, MD).^32^ Data were temporarily stored in a secure database at Copenhagen University Hospital Hvidovre during data collection and subsequently transferred to the University of Copenhagen, Department of Public Health. In addition, serum, plasma and cell fractions from whole blood were extracted and are stored in a biobank at Copenhagen University Hospital Hvidovre for future research.

### Register follow-up

All Danish residents are given a unique 10-digit Civil Personal Registration (CPR) number at birth or upon immigration. This unique CPR number allows for linkage between various type of data, including data from nationwide registers for research purposes. The CPR number was available for all participants in the Population Sample and the Clinical Sample and most of the participants in the 2^nd^ follow-up of the Citizen Science Sample also reported their CPR number. This allowed us to link information on 7,208 individuals with social and health registries in a pseudonymous way. The data linkage includes the Civil Registration System, the Danish Psychiatric Central Register,^33^ the Danish National Prescription Registry,^34^ the Danish Medical Birth Registry,^35^ the Danish in Vitro Fertilization Register,^36^ the Danish National Patient Registry,^37^ Danish Register of Causes of Death,^38^ and various social registers. This linkage will allow us to investigate long-term health and social consequences of night-time smartphone use.

### Patient and public involvement

The public was involved in the initiation of the *SmartSleep Study* through citizen science approaches applied in the baseline data collection of the Citizen Science Sample, including a direct feedback and engagement with the preliminary results.

### Ethical and data approvals

All data collections in the *SmartSleep Study* have been approved under the University of Copenhagen’s umbrella approval from the Danish Data Protection Agency (datatilsynet.dk), which ensures compliance with national and EU legislation in terms of data security (Citizen Science Sample no. 514-0237/18-3000 and 514-0288/19-3000 (2^nd^ follow-up), Population sample no. 514-0288/19-3000 and Clinical Sample no. 514-0344/19-3000). The data collection for the Clinical Sample was further approved by the National Committee on Health Research Ethics (approval no. 67074). Participation in all parts of the study was voluntary and the participants gave informed consent.

## FINDINGS TO DATE

The *SmartSleep Study* is a newly established dataset and only few studies have been published to date,^21,22,39^ but several studies related to mental health, fertility, obesity, and inflammatory profiles are currently ongoing.

### Citizen science as a platform for public health interventions

Citizen science can be defined as research in which the public is engaged in one or more of the research processes.^40^ This approach was used in the Citizen Science Sample of the *SmartSleep Study*, as outlined above. Citizen science has also been highlighted as a strategy to maximize the social impact of research,^41^ and using data from the 1^st^ follow-up of the Citizen Science Sample, we aimed at evaluating whether the massive public focus on sleep and smartphones during the radio campaign influenced night-time smartphone behavior among the participants.^22^ We found that 15% of the participants had changed their night-time smartphone behavior two weeks after the campaign, and that the vast majority of those modifying their behavior (83%) had reduced their night-time use. Increased knowledge and awareness appeared to be the key motivational drivers for these changes. We conclude that public health projects may benefit from exploring citizen science in combination with other interventional approaches.^22^

### Night-time smartphone use and sleep

Night-time smartphone use is highly prevalent in all three samples, especially among young adults (Figure 2). The highest prevalence is found in the youngest age group, where 28% of those aged 16-25 in the Citizen Science Sample used their phone during the sleep period on a regular basis (a few nights a week or more). The corresponding number was 34% among those aged 18-25 in the Population Sample, and 53% among those aged 18-20 in the Clinical Sample. We have previously used the Citizen Science arm of the *SmartSleep Study* to disentangle the effects of day and night-time smartphone use, and we have shown that night-time smartphone use, rather than daytime smartphone, is associated with poorer sleep quality and a higher risk of both short and long sleep duration compared to those who do not use their phone during the sleep period.^21^

**Figure 2.**
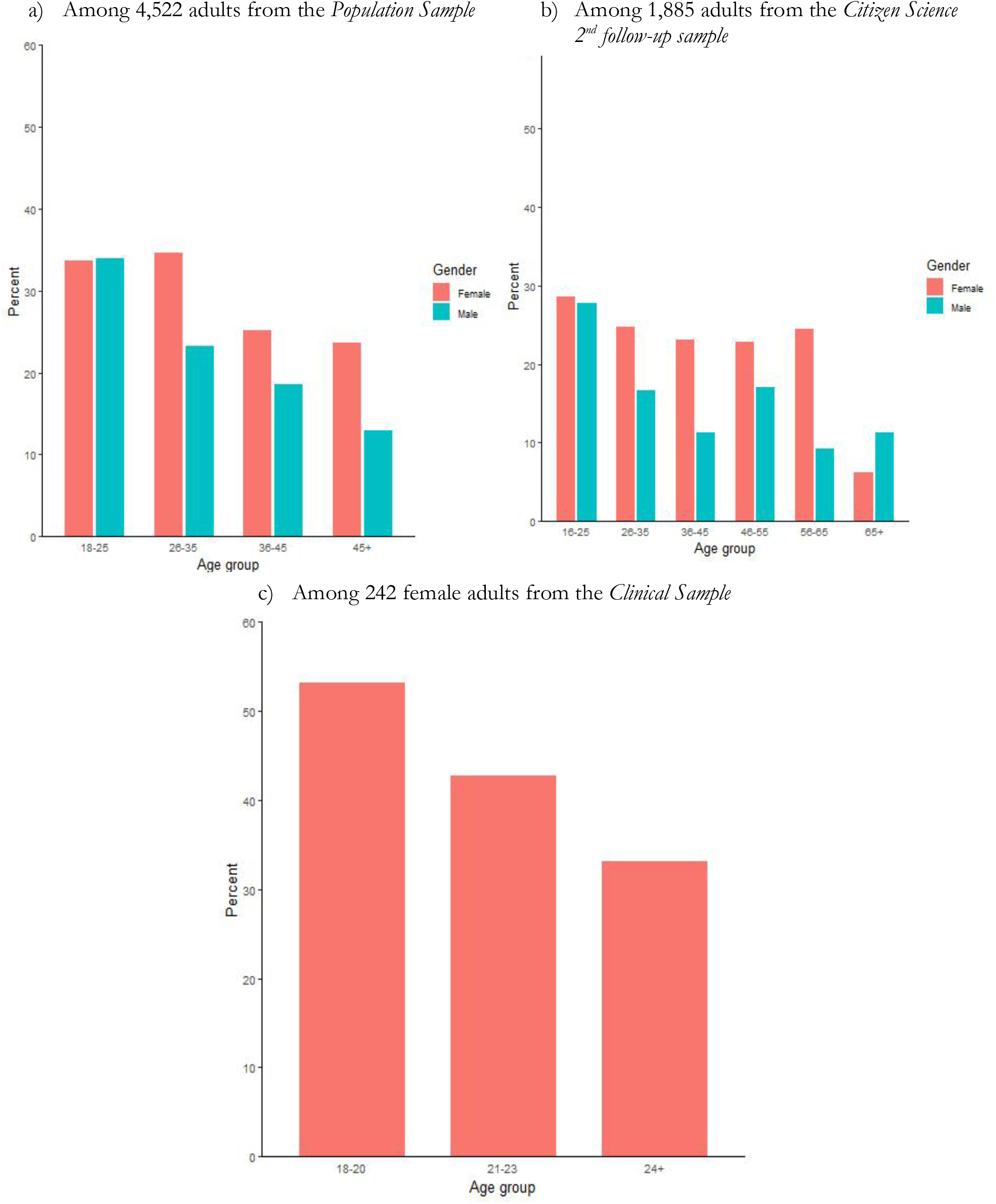
Self-reported night-time smartphone use *a few nights a week or more* according to age and gender.

### Patterns of night-time smartphone use

The *SmartSleep Study* is one of the largest studies so far to have developed an application and collected objective tracking data on smartphone use during night-time over repeated nights. We collected 3.97 million data points in 5,927 individuals via the SmartSleep app. One data point represents either an activation or a de-activation of the phone throughout the data collection period (including both days and nights). In a subsample of 40,625 fully tracked nights among these 5,927 individuals, we present results obtained using non-parametric Functional Data Analysis,^42^ in which we utilize 803,000 data-points from the high-resolution data to identify clusters of temporal trajectories of night-time smartphone use that characterize distinct use patterns. Please see Supplementary Text 2 for an outline of the method and the technical details of the statistical analysis.

Based on the clustering of the temporal activity patterns represented as functional data, we found that four clusters provided the optimal representation of smartphone use according to the optimization criterion. Figure 3 shows the average standardized activity patterns for each of these clusters. The shapes of the clusters show distinct types of night-time smartphone use. The *non-use* cluster includes 37.6% of the nights, and this cluster is characterized by no smartphone use during the sleep period.

**Figure 3.**
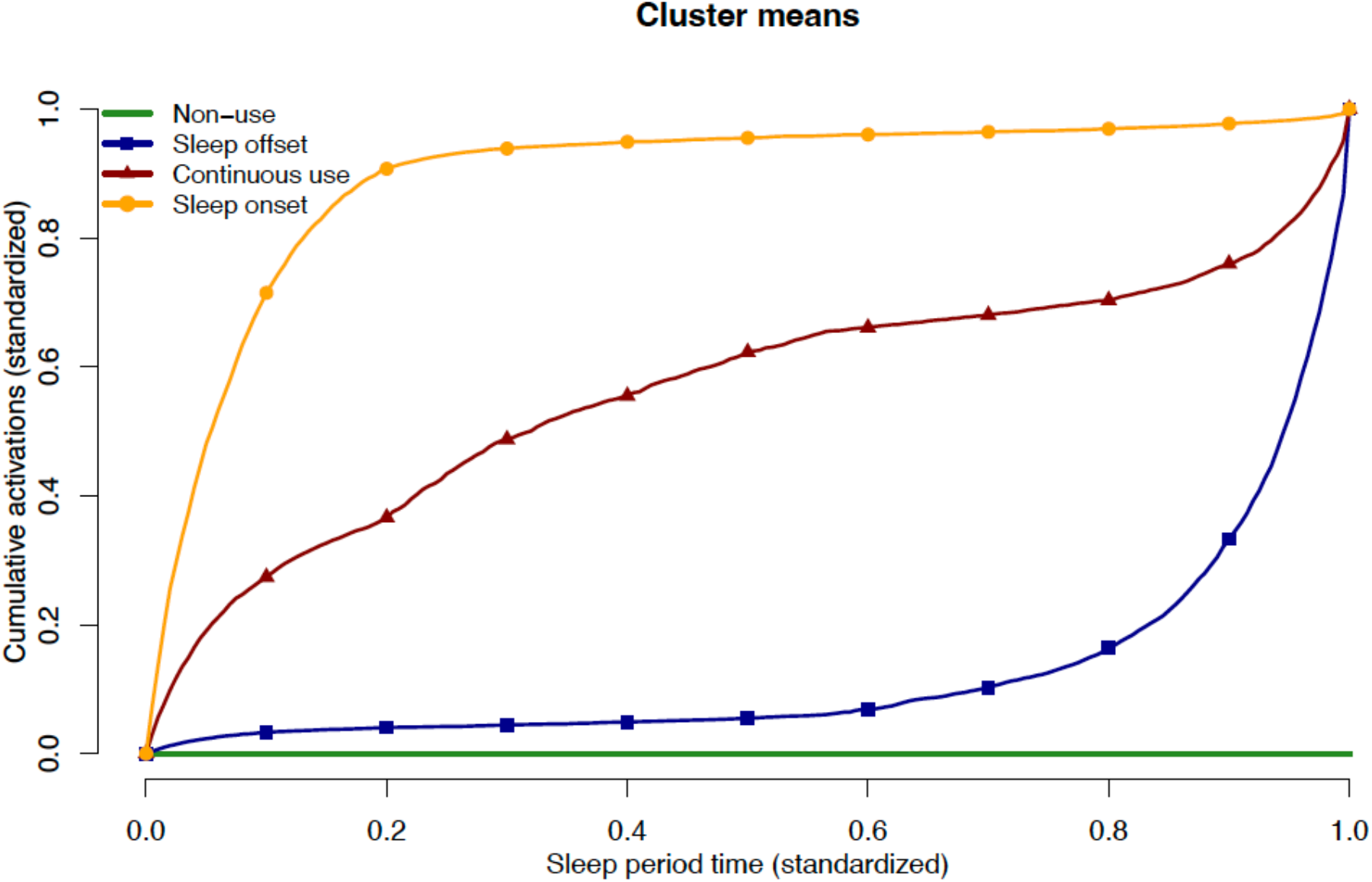
Mean functions of the standardized cumulative nighttime screen activation patterns for each of the four identified clusters. The time scale represents the night-time sleep period (standardized from 0 to 1 for all nights).

The *sleep onset* cluster includes 21.7 % of the nights, and this cluster is characterized by smartphone use during the sleep period, but mainly confined to the period right after sleep onset. The *sleep offset* cluster includes 29.1% of the nights, and this cluster is characterized by smartphone use during the sleep period, but mainly confined to the period right before sleep offset. Finally, 11.6% of the tracked nights belongs to the *continuous use* cluster, which is characterized by repeated smartphone use during the sleep period. In this cluster, the rate of activity increases in the first and last approximately 12.5% of the nightly sleep period as seen by the curvature, but in the middle 75% the slope is close to linear which indicates uniformly distributed activity on average during this period.

Figure 4 shows summary statistics of key features characterizing smartphone use in each of the estimated clusters. In general, the non-use cluster has shorter sleeping periods than the three other clusters. The continuous use and the sleep offset use clusters are associated with later sleep offset times than the other two clusters. The continuous use cluster generally has a higher number of activations and a longer total duration of screen activation during the sleep period than the other clusters.

**Figure 4.**
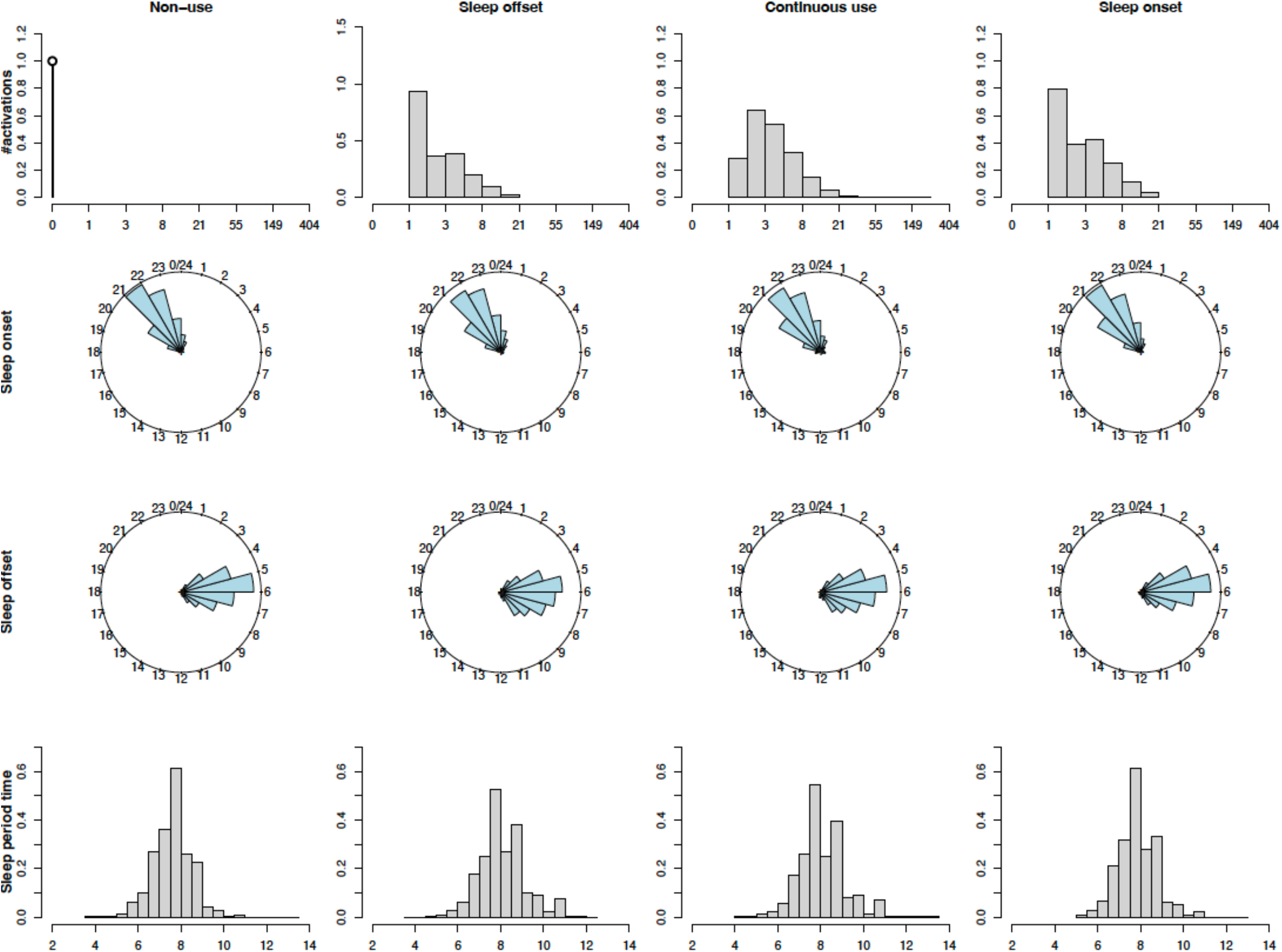
Cluster specific distributions of number of activations, sleep onset and off times and sleep duration.

## STRENGTHS AND LIMITATIONS

The unprecedented size and coverage of the *SmartSleep Study* allow for a comprehensive documentation of smartphone activity during the entire sleep period. The data in the *SmartSleep Study* are sampled in three different ways, which allow us to robustly validate findings across samples. This aligns with the principles of triangulation, which aims at obtaining more reliable answers to research questions through the integration of results from different approaches.^43^ While we may envision a randomized controlled trial aimed at smartphone intervention in a smaller selected sample, it would be difficult – or even impossible – to design a trial which would uncover the day-to-day dynamics of smartphone use, sleep and health in the general population, which is really the public health question of concern. By combining different approaches with different sources of bias, triangulation allows us to address this complex question from different angles.

Table 2 shows an overview of the key sources of bias in each arm of the *SmartSleep Study*. Selection bias due to selective entry into the study is a concern in all three samples, but due to the varying modes of enrollment and data collection in the three samples, we will be able to strengthen the evidence by cross-context comparisons.^43^ The Citizen Science Sample was collected through nationwide radio, which ensured public engagement and a large sample, but it came at the cost of a selected sample without a transparent sampling strategy. Not surprisingly, given the platform for data collection, the participants were more likely to be women, have high education and be younger than the general population.^21^ While we cannot fully document the bias arising from this self-selection, we are most concerned that those with sleep problems were more likely to participate in the study. To address this concern, we have previously compared the prevalence of poor sleep in the Citizen Science Sample with the prevalence in the representative Danish National Health Survey 2017, and it is reassuring to note that we found very similar numbers.^21^

**Table 2.** Sources of bias in the *SmartSleep Study*

|  |  | Bias type |  |  |  |  |
| --- | --- | --- | --- | --- | --- | --- |
|  |  | Selection bias |  | Confounding | Misclassification |  |
|  |  | Selection into study | Loss to follow-up |  | Night-time smartphone use | Health outcomes |
| Cross context comparison | **Citizen Science Sample** | Self-selection, 16 to 99 yrs, men and women, all regions (response proportion unknown) | 72% participated in 1<sup>st</sup> follow-up<br>17% participated in 2<sup>nd</sup> follow-up | Negative control (day-time smartphone use) | Self-reported (survey data) | Self-reported (survey data) |
|  | **Population Sample** | Random sample, 18 to 50 yrs, men and women, all regions (6% response proportion) | Registry linkage (>95% follow-up) | Negative control (day-time smartphone use) | Objectively measured (tracking data) | Objective measured (registry linkage data) |
|  | **Clinical Sample** | Random sample, 18 to 25 yrs, women, capital region (11% response proportion) | Registry linkage (>95% follow-up) | Negative control (day-time smartphone use) | Objectively measured (tracking data) | Objective measured (registry linkage data) + clinical examination |

The Population Sample and the Clinical Sample are randomly drawn from the general population to ensure representativeness, but they are both challenged by relatively low levels of participation. We asked people to fill in a detailed survey and to download the *SmartSleep* app to track their smartphone use, preferably up to 14 nights, in both samples. This is a rather demanding task, and we had initially expected a level of participation similar to those obtained in biobanks. Thus, we are not surprised by the relatively low level of participation of 6 to 17%, compared to e.g. the 5.5% obtained in the UK Biobank.^44^ But the issue of representativeness remains, and similar to the Citizen Science Sample we have compared the age, sex, residential area and educational distribution in each sample with that of the general population, and we find that more females, middle-aged people, and individuals with a higher educational level participate in the study (Supplementary Table 1). Also, there is an overrepresentation of people living in the Capital Region in the Citizen Science Sample. The level of sleep problems seems similar that of the general population. In the Population Sample, 40% of the population reported disturbed sleep at least a few times a week, and of that number, 9% reported disturbed sleep every night or almost every night. In the Danish National Health Survey 2021, 49% of the 173,324 participants reported a few or several sleep impairments within the past 14 days, of which 15% had several sleep impairments.^45^

Loss to follow-up is another potential source of selection bias which may impact the results. The *SmartSleep Study* benefits from applying different means of follow-up, which allow us to triangulate evidence across samples with varying degree of loss to follow-up. A strength of the *SmartSleep Study* is that a large subsample of the study population (n=7,208) has agreed to be prospectively followed-up in nationwide registers in years to come for medication use, mental and physical health diagnoses and social outcomes through the personal identification number assigned to every Danish citizen at birth. This allows for almost complete long-term follow-up for key health outcomes.

Confounding is an inevitable source of bias in observational studies, including the *SmartSleep Study*. Those who report high levels of smartphone use may differ from those who do not, e.g., by age and gender. Some of these differences can be adjusted for in our statistical analyses, but residual confounding from unmeasured or poorly measured confounders will always remain a concern. One way to address such concerns is to identify so-called ‘negative control measures’, which are characterized by sharing the confounder structure of the exposure of interest while ideally being unrelated to the outcome.^43^ The *SmartSleep Study* includes comprehensive measures of day-time smartphone use, which most likely share the same confounding structure as night-time smartphone use, but is hypothesized to be unrelated (or at least less related) to the outcomes of interest, which are primarily hypothesized to be impacted through mechanisms of impaired sleep (e.g. fertility and immune system functioning). This is supported by our initial findings, where day-time smartphone use was only weakly associated with sleep impairment, when night-time use was considered.^21^

Misclassification is also a source of systematic bias in epidemiological studies. The *SmartSleep Study* benefits from applying different approaches to data collection, which will allow us to triangulate evidence across samples with potentially different and varying sources of misclassification. Most previous studies solely relied on survey questions, but in addition to comprehensive survey data we also developed an app to objectively track night-time smartphone use. This allowed us to perform an unprecedented characterization of patterns of night-time smartphone use without problems with recall or social desirability bias. Also, by clustering of the activity shape functions of smartphone use via Functional Data Analysis we obtained a collection of salient characteristics representing different dynamical properties of this use. Another potential source of misclassification relates to the lack of objective sleep assessment in the study. Subjective sleep reports tend to overestimate time in bed and sleep time,^46,47^ and it is therefore likely that in some cases our tracking results are not strictly limited to the sleep period but might also include short wake periods before sleep onset and after awakening. While this seems negligible for most research questions, we suggest not to use our dataset to investigate research questions that require an unambiguous (objective) sleep onset time point.

In conclusion, the three samples nested within the *SmartSleep Study* represent multi-dimensional, temporal data, which allow for triangulation of evidence from different sources of evidence (surveys, tracking, clinical, and registry data) with varying degrees of bias. This design is expected to provide a reliable evidence base for the study of the dynamic interplay between night-time smartphone use, sleep and health.

## Data Availability

The dataset contains personal identifiable data and sensitive information. We are therefore not allowed to make them publicly available according to the Danish Protection Agency (Danish data protection legislation datatilsynet.dk) and Danish law. Inquiries for secure access under conditions stipulated by the Danish Data Protection Agency should be directed at the principal investigator of the SmartSleep Study Professor Naja Hulvej Rod.

## Collaboration

We encourage collaboration, and access to the *SmartSleep Study* is available to other investigators through collaborative agreements and a secured access. Please contact Professor Naja Hulvej Rod for further information.

## Data sharing statement

The dataset contains personal identifiable data and sensitive information. We are therefore not allowed to make them publicly available according to the Danish Protection Agency (Danish data protection legislation datatilsynet.dk) and Danish law. Inquiries for secure access under conditions stipulated by the Danish Data Protection Agency should be directed at the principal investigator of the *SmartSleep Study* Professor Naja Hulvej Rod.

## Funding

The *SmartSleep Study* is funded by a Sapere Aude grant from the Independent Research Fund Denmark (grant number 7025-00005B), Helsefonden (grant number 20-B-0254) and the Velliv Association (grant number grant number 20-0047). The funders had no role in study design, data collection and analysis, decision to publish, or preparation of the manuscript.

## Contributors

The *SmartSleep Study* was conceptualized by NH Rod, AS Dissing, TO Andersen, HS Nielsen, NlC Freiesleben and ER Severinsen and further developed by AK Jensen, TV Varga and HJ Drews. The data was cleaned and analyzed by TO Andersen, TV Varga, VT Pham, C Sejling, LKH Schmidt, and AK Jensen. Data were validated in a subsample by M Nygaard, TO Andersen and C Sejling. NH Rod wrote the first draft of the paper. All authors were involved in the revisions of the paper and approved the submitted version. AS Dissing’s contribution to the paper was confined to her previous position as Assistant Professor at University of Copenhagen, and not her current position at Lundbeck A/S.

## Competing interests

No competing interests.

## Notes

### Competing Interest Statement

The authors have declared no competing interest.

### Author Declarations

All data collections in the SmartSleep Study have been approved under the University of Copenhagen umbrella approval from the Danish Data Protection Agency (datatilsynet.dk), which ensures compliance with national and EU legislation in terms of data security (Citizen Science Sample no. 514-0237/18-3000 and 514-0288/19-3000 (2nd follow-up), Population sample no. 514-0288/19-3000 and Clinical Sample no. 514-0344/19-3000). The data collection for the Clinical Sample was further approved by the National Committee on Health Research Ethics (approval no. 67074). Participation in all parts of the study was voluntary and the participants gave informed consent.

## REFERENCES

1 Joiner WJ. Unraveling the Evolutionary Determinants of Sleep. Current Biology 2016; 26: R1073–87.

2 Medic G, Wille M, Hemels MEH. Short- and long-term health consequences of sleep disruption. Nature and Science of Sleep 2017; 9: 151.

3 Wood AW, Loughran SP, Stough C. Does evening exposure to mobile phone radiation affect subsequent melatonin production? International Journal of Radiation Biology 2006. DOI:10.1080/09553000600599775.

4 Loughran SP, Wood AW, Barton JM, Croft RJ, Thompson B, Stough C. The effect of electromagnetic fields emitted by mobile phones on human sleep. NeuroReport 2005; 16: 1973–6.

5 Schmid SR, Höhn C, Bothe K, et al. How Smart Is It to Go to Bed with the Phone? The Impact of Short-Wavelength Light and Affective States on Sleep and Circadian Rhythms. Clocks & Sleep 2021, Vol 3, Pages 558-580 2021; 3: 558–80.

6 Pandi-Perumal SR, Monti JM, Burman D, et al. Clarifying the role of sleep in depression: A narrative review. Psychiatry Research. 2020; 291. DOI:10.1016/j.psychres.2020.113239.

7 Riemann D, Berger M, Voderholzer U. Sleep and depression - Results from psychobiological studies: An overview. Biological Psychology 2001; 57: 67–103.

8 Li Y, Li G, Liu L, Wu H. Correlations between mobile phone addiction and anxiety, depression, impulsivity, and poor sleep quality among college students: A systematic review and meta-analysis. Journal of Behavioral Addictions. 2020; 9: 551–71.

9 Thomée S. Mobile Phone Use and Mental Health. A Review of the Research That Takes a Psychological Perspective on Exposure. 2018. DOI:10.3390/ijerph15122692.

10 Jørgensen N, Joensen UN, Jensen TK, et al. Human semen quality in the new millennium: A prospective cross-sectional population-based study of 4867 men. BMJ Open 2012; 2. DOI:10.1136/bmjopen-2012-000990.

11 Leproult R, Van Cauter E. Effect of 1 week of sleep restriction on testosterone levels in young healthy men. JAMA - Journal of the American Medical Association. 2011; 305: 2173– 4.

12 Jensen TK, Andersson AM, Skakkebæk NE, et al. Association of sleep disturbances with reduced semen quality: A cross-sectional study among 953 healthy young Danish men. American Journal of Epidemiology 2013; 177: 1027–37.

13 Exelmans L, Van den Bulck J. Bedtime mobile phone use and sleep in adults. Social Science & Medicine 2016; 148: 93–101.

14 Carter B, Rees P, Hale L, Bhattacharjee D, Paradkar MS. Association Between Portable Screen-Based Media Device Access or Use and Sleep Outcomes: A Systematic Review and Meta-analysis. JAMA Pediatr 2016; 170: 1202–8.

15 Vernon L, Modecki KL, Barber BL. Mobile Phones in the Bedroom: Trajectories of Sleep Habits and Subsequent Adolescent Psychosocial Development. 2017. DOI:10.1111/cdev.12836.

16 Rod NH, Dissing AS, Clark A, Gerds TA, Lund R. Overnight smartphone use: A new public health challenge? A novel study design based on high-resolution smartphone data. PLoS ONE 2018; 13. DOI:10.1371/journal.pone.0204811.

17 Christensen MA, Bettencourt L, Kaye L, et al. Direct Measurements of Smartphone Screen-Time: Relationships with Demographics and Sleep. PLoS One 2016; 11. DOI:10.1371/JOURNAL.PONE.0165331.

18 BBC Radio 4 - The Anatomy of Loneliness - Who feels lonely? The results of the world’s largest loneliness study. https://www.bbc.co.uk/programmes/articles/2yzhfv4DvqVp5nZyxBD8G23/who-feels-lonely-the-results-of-the-world-s-largest-loneliness-study (accessed Aug 16, 2021).

19 Hammond C, Lewis G. The Rest Test: Preliminary Findings from a Large-Scale International Survey on Rest. In: The Restless Compendium. Springer International Publishing, 2016: 59– 67.

20 Den Broeder L, Devilee J, Van Oers H, Schuit AJ, Wagemakers A. Citizen Science for public health. Health Promot Int 2018. DOI:10.1093/heapro/daw086.

21 Dissing AS, Andersen TO, Nørup LN, Clark A, Nejsum M, Rod NH. Daytime and nighttime smartphone use: A study of associations between multidimensional smartphone behaviours and sleep among 24,856 Danish adults. Journal of Sleep Research 2021; : e13356.

22 Andersen TO, Dissing AS, Varga T V., Rod NH. The SmartSleep Experiment: Evaluation of changes in night-time smartphone behavior following a mass media citizen science campaign. PLOS ONE 2021; 16: e0253783.

23 Priskorn L, Nordkap L, Bang AK, et al. Average sperm count remains unchanged despite reduction in maternal smoking: results from a large cross-sectional study with annual investigations over 21 years. Hum Reprod 2018; 33: 998–1008.

24 Lund R, Nielsen LS, Henriksen PW, Schmidt L, Avlund K, Christensen U. Content validity and reliability of the copenhagen social relations questionnaire. Journal of Aging and Health 2014; 26: 128–50.

25 Nordin M, Åkerstedt T, Nordin S. Psychometric evaluation and normative data for the karolinska sleep questionnaire. Sleep and Biological Rhythms 2013; 11: 216–26.

26 Cohen S, Kamarck T, Mermelstein R. A global measure of perceived stress. J Health Soc Behav 1983.

27 Eskildsen A, Dalgaard VL, Nielsen KJ, et al. Cross-cultural adaptation and validation of the danish consensus version of the 10-item perceived stress scale. Scandinavian Journal of Work, Environment and Health 2015; 41: 486–90.

28 Bech P, Rasmussen NA, Olsen LR, Noerholm V, Abildgaard W. The sensitivity and specificity of the Major Depression Inventory, using the Present State Examination as the index of diagnostic validity. Journal of Affective Disorders 2001; 66: 159–64.

29 Diener E, Emmons RA, Larsem RJ, Griffin S. The Satisfaction With Life Scale. Journal of Personality Assessment 1985; 49: 71–5.

30 Hughes ME, Waite LJ, Hawkley LC, Cacioppo JT. A short scale for measuring loneliness in large surveys: Results from two population-based studies. Research on Aging. 2004; 26: 655–72.

31 Billieux L, Van Der Linden M, Rochat L. The Role of Impulsivity in Actual and Problematic Use of the Mobile Phone. DOI:10.1002/acp.1429.

32 V-PLEX Human Biomarker 54-Plex Kit | Meso Scale Discovery. https://www.mesoscale.com/products/v-plex-human-biomarker-54-plex-kit-k15248d/ (accessed March 3, 2022).

33 Mors O, Perto GP, Mortensen PB. The Danish psychiatric central research register. Scandinavian Journal of Public Health 2011. DOI:10.1177/1403494810395825.

34 Pottegård A, Schmidt SAJ, Wallach-Kildemoes H, Sørensen HT, Hallas J, Schmidt M. Data Resource Profile: The Danish National Prescription Registry. Int J Epidemiol 2017; 46: 798.

35 Bliddal M, Broe A, Pottegård A, Olsen J, Langhoff-Roos J. The Danish Medical Birth Register. European Journal of Epidemiology; 33. DOI:10.1007/s10654-018-0356-1.

36 Andersen AN, Erb K. Register data on assisted reproductive technology (ART) in Europe including a detailed description of ART in Denmark. DOI:10.1111/j.1365-2605.2005.00577.x.

37 Schmidt M, Schmidt SAJ, Sandegaard JL, Ehrenstein V, Pedersen L, Sørensen HT. The Danish National patient registry: A review of content, data quality, and research potential. Clinical Epidemiology. 2015. DOI:10.2147/CLEP.S91125.

38 Helweg-Larsen K. The Danish Register of Causes of Death. Scand J Public Health 2011; 39: 26–9.

39 Andersen TO, Dissing AS, Severinsen ER, et al. Predicting Stress and Depressive Symptoms Using High-Resolution Smartphone Data and Sleep Behaviour in Danish Adults. Sleep 2022; published online March 17. DOI:10.1093/SLEEP/ZSAC067.

40 Phillips T, Porticella N, Constas M, Bonney R. A Framework for Articulating and Measuring Individual Learning Outcomes from Participation in Citizen Science. Citizen Science: Theory and Practice 2018; 3: 3.

41 Brouwer S, Hessels LK. Increasing research impact with citizen science: The influence of recruitment strategies on sample diversity: https://doi.org/101177/0963662519840934 2019; 28: 606–21.

42 Wang J-L, Chiou J-M, Müller H-G. Functional Data Analysis. Annual Review of Statistics and Its Application 2016; 3: 257–95.

43 DA L, K T G DS. Triangulation in aetiological epidemiology. Int J Epidemiol 2016; 45: 1866–86.

44 Fry A, Littlejohns TJ, Sudlow C, et al. Comparison of Sociodemographic and Health-Related Characteristics of UK Biobank Participants With Those of the General Population. American Journal of Epidemiology 2017; 186: 1026–34.

45 Sundhedsstyrelsen. Danskernes sundhed – Den Nationale Sundhedsprofil 2021. Rosendahls A/S, 2022.

46 Lehrer HM, Yao Z, Krafty RT, et al. Comparing polysomnography, actigraphy, and sleep diary in the home environment: The Study of Women’s Health Across the Nation (SWAN) Sleep Study. SLEEP Advances 2022; 3. DOI:10.1093/SLEEPADVANCES/ZPAC001.

47 Girschik J, Fritschi L, Heyworth J, Waters F. Validation of Self-Reported Sleep Against Actigraphy. Journal of Epidemiology 2012; 22: 462–8.

48 Ward Jr JH. Hierarchical grouping to optimize an objective function. Journal of the American statistical association 1963; 58.301: 236–44.

49 Charrad M, Ghazzali N, Boiteau V, Niknafs A. NbClust: An R Package for Determining the Relevant Number of Clusters in a Data Set. Journal of Statistical Software 2014; 61: 1–36.

